# Safety and immunogenicity of the BBIBP-CorV vaccine in adolescents aged 12-17 years in Thai population, prospective cohort study

**DOI:** 10.1101/2022.01.07.22268883

**Authors:** Kriangkrai Tawinprai, Taweegrit Siripongboonsitti, Thachanun Porntharukchareon, Preeda Vanichsetakul, Saraiorn Thonginnetra, Krongkwan Niemsorn, Pathariya Promsena, Manunya Tandhansakul, Naruporn Kasemlawan, Natthanan Ruangkijpaisal, Narin Banomyong, Nanthida Phattraprayoon, Teerapat Ungtrakul, Kasiruck Wittayasak, Nawarat Thonwirak, Kamonwan Soonklang, Gaidganok Sornsamdang, Nithi Mahanonda

**Affiliations:** Department of Medicine, Chulabhorn Hospital, Chulabhorn Royal Academy; Department of Pediatrics, Chulabhorn Hospital, Chulabhorn Royal Academy; Princess Srisavangavadhana College of Medicine, Chulabhorn Royal Academy; Center of Learning and Research in Celebration of HRH Princess Chulabhorn’s 60th Birthday, Chulabhorn Royal Academy; Central Laboratory Center, Chulabhorn Hospital, Chulabhorn Royal Academy

**Author notes:** Corresponding author: Kriangkrai Tawinprai, 906 Kamphaeng Phet 6 Rd., Talat Bang Khen, Lak Si, Bangkok 10210 email.

**Keywords:** COVID-19, SARS-CoV-2, Adolescent, BBIBP-CorV, Safety, Immunogenicity

## Abstract

**Introduction:** COVID-19 pandemic affects all populations worldwide, including adolescents. Adolescents can develop a severe form of COVID-19, especially with comorbidity underlying. The prior studies of the mRNA COVID-19 vaccine showed excellent effectiveness in adolescents. Therefore, this study aimed to evaluate the safety and effectiveness of the BBIBP-CorV vaccine with the immunobridging approach in Thai adolescents.

**Methods:** This single-center, prospective cohort study compared the immunogenicity after 2 doses of the BBIBO-CorV vaccine with 21 days interval of participants aged 12-17 years with 18-30 years at Chulabhorn Hospital, Chulabhorn Royal Academy, Bangkok, Thailand. The key eligible criteria were healthy or had stable pre-existing comorbidity participants, aged 12-17 years. The primary endpoint was the anti-receptor binding domain antibody concentration at 4 weeks after dose 2 of the vaccine. In addition, safety profiles were solicited adverse events within 7 days after each dose of vaccine and any adverse events through 1 month after dose 2 of the vaccine.

**Results:** Four weeks after the second vaccination, the GMC of anti-RBD antibody in the adolescent cohort was 102.9 BAU/mL (95%CI; 91.0-116.4) and 36.9 BAU/mL (95%CI; 30.9-44.0) in the adult cohort. The GMR of the adolescent cohort was 2.79 (95%CI; 2.25-3.46, p-value; <0.0001) compared with the adult cohort which met non-inferiority criteria. The reactogenicity was slightly less reported in the adolescent cohort compared with the adult cohort. No serious adverse events were reported in both cohorts.

**Conclusion:** Vaccination with the BBIBP-CorV vaccine in the adolescent participants was safe and effective.

## INTRODUCTION

COVID-19 pandemic affects all populations worldwide, including adolescents. In general, most COVID-19 children or adolescents were a mild form of severity. (1) However, some adolescents can develop a severe form of COVID-19, especially with comorbidity underlying. (2) In addition, there were reports of a complication related to a severe acute respiratory syndrome coronavirus 2 (SARS-CoV-2) in children and adolescents, a multisystem inflammatory syndrome in children (MIS-C). (3) MIS-C is a syndrome related to SARS-CoV-2 infection. Multiorgan characteristics, especially cardiovascular and mucocutaneous, involvement, and extreme inflammation. (4)

Prior studies in adults showed that the COVID-19 vaccines could prevent symptomatic SARS-CoV-2 infection, hospitalization, and death. (5-8) However, the studies of the safety and effectiveness of the COVID-19 vaccine in children and adolescents were limited. There are two studies of the BNT 162b2 vaccine in children aged 5-11 years and 12-15 years. These studies showed a favorable safety profile and immunogenicity compared with the adult cohort. (9, 10)

The BBIBP-CorV vaccine is the inactivated COVID-19 vaccine developed by the Beijing Bio-Institute of Biological Products, China. The phase 3 clinical trial demonstrated efficacy and safety, including participants aged 18 years or more. The efficacy for preventing symptomatic COVID-19 was high, up to 78.1% (8). The safety profile after vaccination in children and adolescents aged 3-17 years in phase 1/2 trials was favorable. (11) However, the study of effectiveness is lacking.

This study aimed to evaluate the safety profile and the effectiveness of the BBIBP-CorV vaccine with the immunobridging approach in Thai adolescents aged 12-17 years.

## METHODS

### Trial design and Participants

We conducted a single-center, prospective cohort study to evaluate the immunogenicity and safety of BBIBP-CorV vaccine in participants aged 12-17 years (adolescent cohort) compared with 18-30 years (adult cohort) at Chulabhorn Hospital, Chulabhorn Royal Academy, Bangkok, Thailand. The key eligible criteria were healthy or had stable pre-existing comorbidity participants, aged 12-17 years. The data of the comparison adult cohort, aged 18-30 years, were randomly selected from the prior safety and immunogenicity study of the BBIBP-CorV vaccine in adult participants in our institute. Participants with known previous SARS-CoV-2 infection, prior COVID-19 vaccination were excluded. All participants and parents were informed consent. The study protocol, informed consent, and case record form were reviewed and approved by the Ethics Committee for Human Research, Chulabhorn Research Institute (reference number: 123/2564). The study was registered with thaiclinicaltrials.org (TCTR20210920005).

### Interventions

All participants were assigned to receive two doses of 4 μg (0.5 mL) of BBIBP-CorV vaccine intramuscularly at 21 days intervals. After each vaccination, all participants were observed immediate adverse events at the clinic for 30 minutes.

### Safety

The local and systemic reactogenicities were evaluated using a questionnaire sent by mobile text message on day-1 and day-7 post-vaccination each dose. In addition, the unsolicited and serious adverse events (AEs) were followed up through 1 month using the mobile text message questionnaire on day-28 post-vaccination and the hotline mobile for participants to inform their adverse events. Pain, tenderness, redness, and swelling were defined as local reactogenicity. On the other hand, fever, nausea or vomiting, diarrhea, headache, fatigue, and myalgia were defined as systemic reactogenicity. Local adverse events were graded as mild if the lesion was less than 5 cm, moderate if the lesion was between 5 to 10 cm, severe if the lesion was more than 10 cm, and life-threatening if the participants needed an emergency department visit or hospitalization. In addition, systemic adverse events were graded as mild if AE did not interfere with daily activity, moderate if AE interfered some daily activity, severe if AE limited daily activity, and life-threatening if the participants needed an emergency department visit or hospitalization.

### Immunogenicity

The immunogenicity was assessed by the anti-receptor-binding domain (RBD) of the spike protein of SARS-CoV-2 concentration, which was performed before the first vaccination and 1 month after the second vaccination using Elecsys Anti-SARS-CoV-2 S (Elecsys-S) kit (Roche Diagnostics, Mannheim, Germany), an automated electrochemiluminescence immunoassay. The test was performed following the manufacture’s instructor. The concentration of the anti-RBD antibody was demonstrated in this study by binding antibody unit per milliliter (BAU/mL). To convert the Elecsys-S unit to BAU, using the Elecsys-S U = 0.972 × BAU equation. (12)

### Outcomes

The primary outcome was the geometric mean of anti-RBD antibody concentration (GMC) at 1 month after the second vaccination between adolescent and adult cohorts. In addition, a comparison between GMC of an anti-RBD antibody of the adolescent cohort and the adult cohort was shown with the geometric mean ratio (GMR) using a non-inferiority fashion.

The secondary outcomes were 1) reactogenicity within 7 days after the BBIBP-CorV vaccination, 2) serious adverse events within 1 month after the second vaccination.

### Sample size

A sample of 250 participants in each cohort was estimated for providing 80% power, 0.05 type I error, and 25% dropout for declaring non-inferiority. (10) Non-inferiority was defined as a lower limit of the two-side 95% confidence interval of geometric mean ratio > 0.67.

### Statistical methods

The anti-RBD antibody’s geometric mean concentration (GMC) was calculated by exponential the arithmetic means of the logarithm of anti-RBD antibody concentration. Comparison between the arithmetic means of the logarithm of anti-RBD antibody concentration between the adolescent and adult cohort using independent-sample t-test. The difference of arithmetic means then was exponential to the geometric mean ratio between the adolescent and adult cohorts. If the non-inferiority criterion was met, then superiority was assessed. Summary statistics were presented as medians and interquartile ranges (IQRs) or geometric mean concentration and 95% confidence intervals (CIs). Statistical analyses were performed using IBM GraphPad Prism version 9 and SPSS statistic version 26. The p-value of less than 0.05 was considered statistically significant.

## RESULTS

### Participants

Between September 14, 2021, and October 5, 2021, a total of 248 adolescent participants were assessed for eligibility criteria. Fourteen participants were excluded. Five participants were excluded due to seropositive status at baseline, and nine could not evaluate baseline serostatus. For the adult cohort, 252 participants aged 18-30 were assessed for eligibility. Three were excluded due to seropositive status at baseline. Finally, 234 participants in the adolescent cohort and 249 in the adult cohort were assigned to receive the study vaccine and were analyzed for safety profile. All participants in the study completed two doses of the study vaccines. However, one month after the second vaccination, 393 (81.4%) followed up for primary outcome analysis, 190 in the adolescent cohort and 203 in the adult cohort. Summarisation of participants’ flow is shown in figure 1.

**Figure 1.**
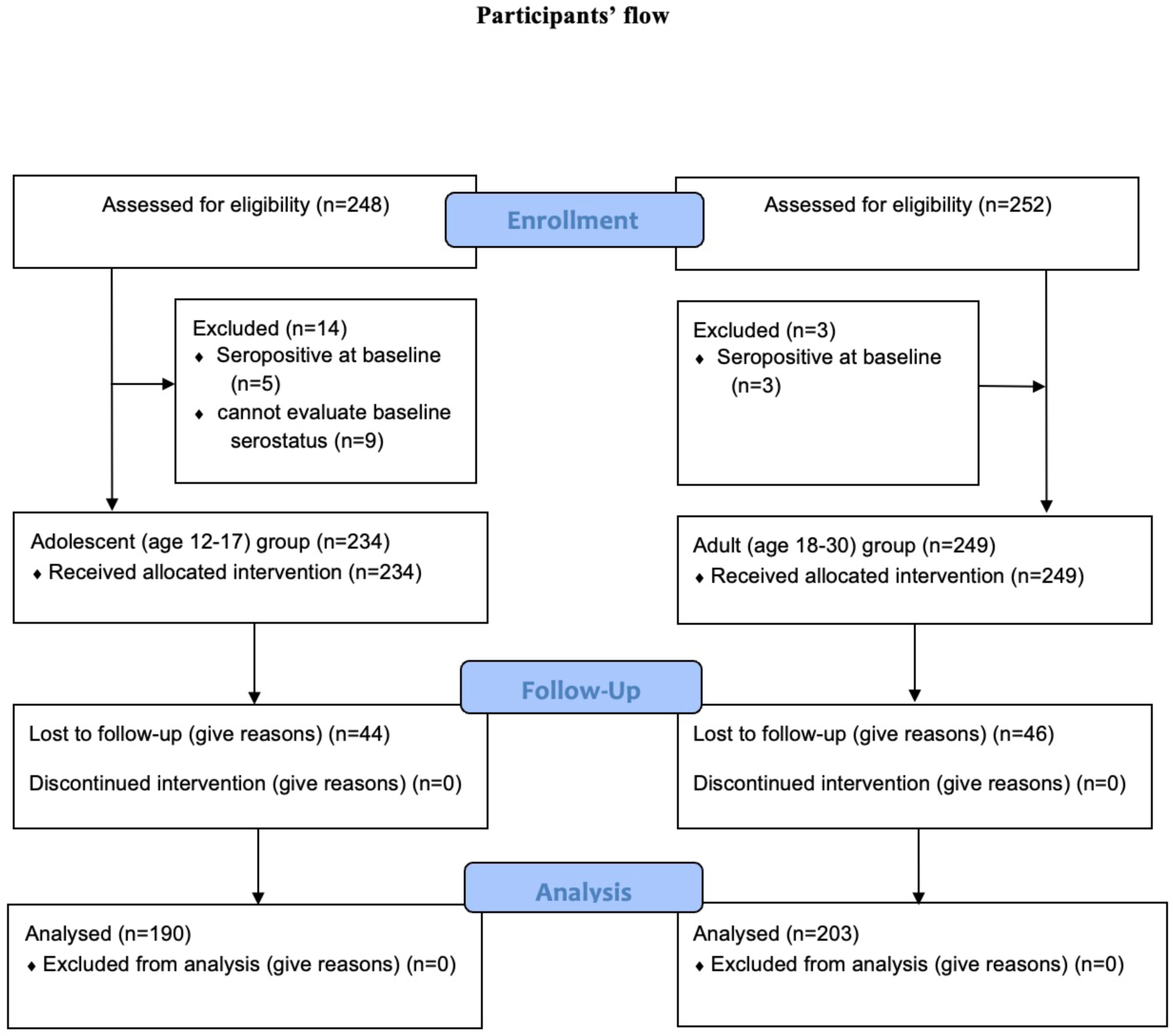
Participants’ flow. Between September 14, 2021, and October 5, 2021, a total of 248 adolescent participants were assessed for eligibility criteria. Fourteen participants were excluded. Five participants were excluded due to seropositive status at baseline, and nine could not evaluate baseline serostatus. Two hundred fifty-two participants aged 18-30 were assessed for eligibility for the adult cohort. Three were excluded due to seropositive status at baseline. Finally, 234 participants in the adolescent cohort and 249 in the adult cohort were assigned to receive the study vaccine and were analyzed for safety profile. One month after the second vaccination, 393 (81.4%) followed up for primary outcome analysis, 190 in the adolescent group and 203 in the adult group.

A baseline demographic data were summarised in table 1. Half of the participants (46.2%) were female in the adolescent cohort, and the median age was 14 (13-16) year-olds. For the adult cohort, 47.4% were female, and the median age was 25 (2-28) year-olds. There was no participant with underlying comorbidity.

**Table 1.** Demographic data of each study group.

|  | 12-17 year | 18-30 year |
| --- | --- | --- |
| Number of participants for safety analysis. | 234 | 249 |
| Female (%). | 108 (46.2%) | 118 (47.4%) |
| Median age (IQR) year. | 14 (13-16) | 25 (22-28) |
| Number of participants for the primary endpoint. | 190 | 203 |
| Female (%). | 90 (47.4%) | 97 (47.8%) |
| Median age (IQR) year. | 14 (13-16) | 25 (23-28) |
| Underlying disease. | 0 | 0 |
| IQR: interquartile range. |  |  |

### Safety

#### Reactogenicity

The summarisations of reactogenicity after the BBIBP-CorV vaccination in adolescent and adult cohorts were demonstrated in figures 2 and 3, respectively. The reactogenicity was slightly less reported in the adolescent cohort compared with the adult cohort. In both cohort studies, the reactogenicity was reported somewhat higher after the first dose than the second dose. On day-1 after the first dose, 18.1% in adolescents and 23.7% in reactogenicity was reported in adults. Most reactions were mild to moderate in severity and resolved within 7 days. One participant in the adolescent cohort reported severe nausea and vomit and spontaneously resolved without a hospital visit.

**Figure 2.**
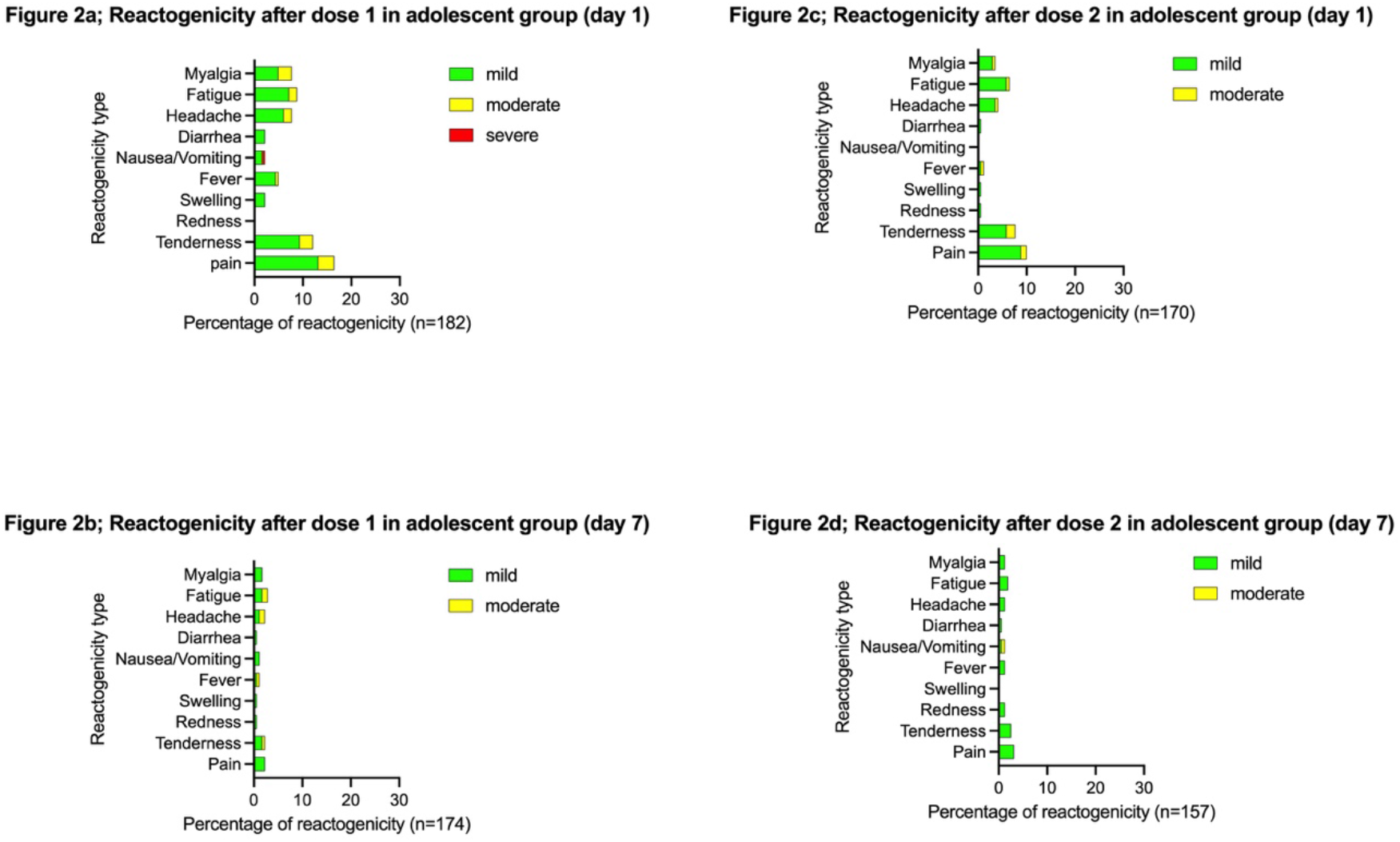
Reactogenicity within 7 days after the BBIBP-CorV vaccination in adolesenct group aged 12-17 years. Pain, tenderness, redness, and swelling were defined as local reactogenicity. Fever, nausea or vomiting, diarrhea, headache, fatigue, and myalgia were defined as systemic reactogenicity. Local adverse events were graded as mild if the lesion was less than 5 cm, moderate if the lesion was between 5 to 10 cm, severe if the lesion was more than 10 cm, and life-threatening if the participants needed an emergency department visit or hospitalization. Systemic adverse events were graded as mild if adverse event did not interfere with daily activity, moderate if adverse event interfered some daily activity, severe if adverse event limited daily activity, and life-threatening if the participants needed an emergency department visit or hospitalization. n; numbers of participants responsed the questionaire.

**Figure 3.**
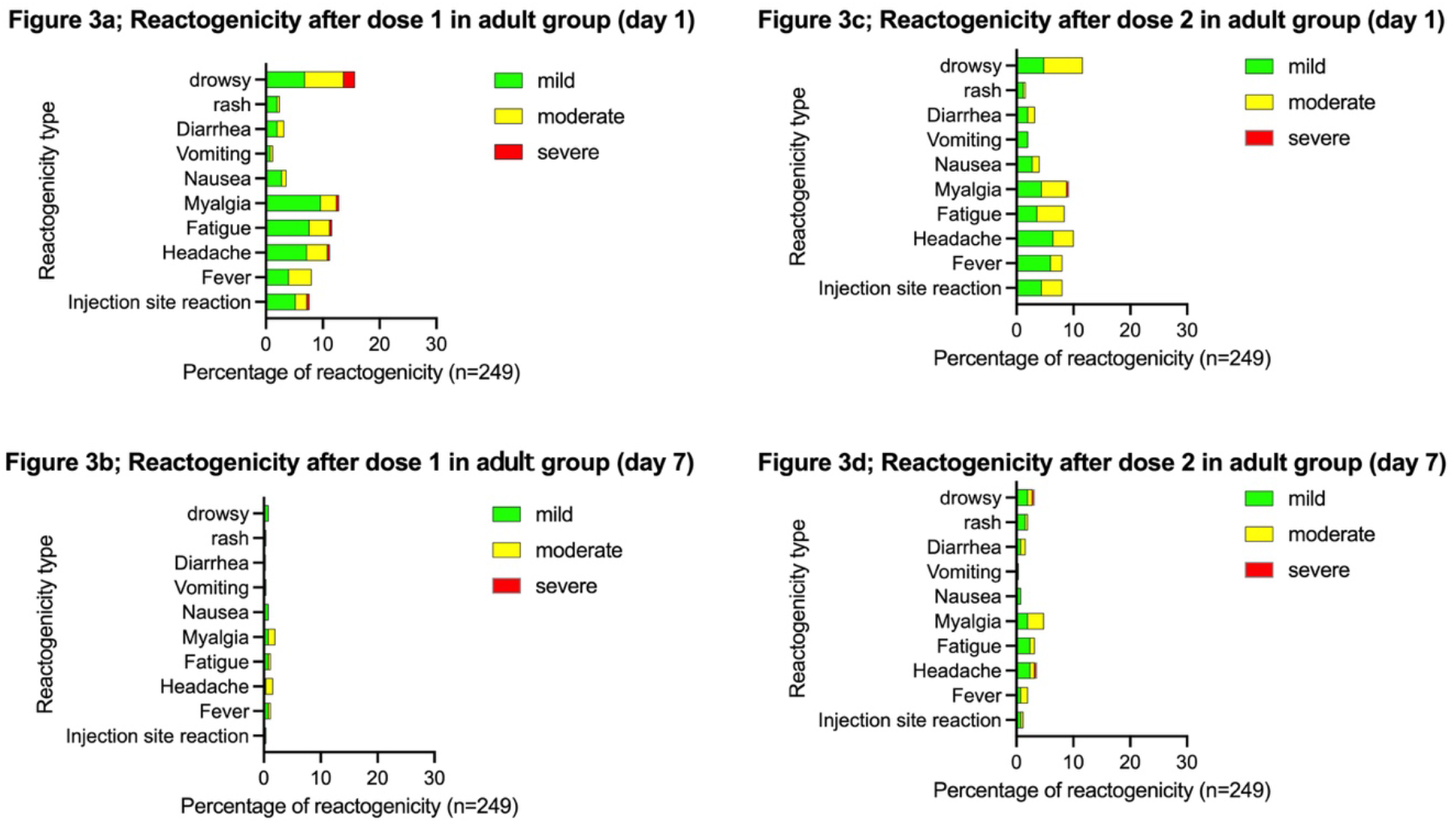
Reactogenicity within 7 days after the BBIBP-CorV vaccination in adult group aged 18-30 years. Local reactogenicity including pain, tenderness, redness, and swelling. Fever, nausea or vomiting, diarrhea, headache, fatigue, and myalgia were defined as systemic reactogenicity. Local adverse events were graded as mild if the lesion was less than 5 cm, moderate if the lesion was between 5 to 10 cm, severe if the lesion was more than 10 cm, and life-threatening if the participants needed an emergency department visit or hospitalization. Systemic adverse events were graded as mild if adverse event did not interfere with daily activity, moderate if adverse event interfered some daily activity, severe if adverse event limited daily activity, and life-threatening if the participants needed an emergency department visit or hospitalization. n; numbers of participants responsed the questionaire.

On the other hand, seven participants in the adult cohort reported nine severe reactogenicities, one injection site reaction, one headache, one fatigue, one myalgia, and five drowsiness. However, they resolved within one week. On day-1 after the second dose, the reactogenicity was reported in 12.4% in an adolescent cohort and 14.1% in the adult cohort. All participants’ reactions were mild to moderate in severity except one in the adult cohort who reported severe myalgia, which spontaneously resolved. There was no serious adverse event reported in the follow-up period.

Local reactions were reported more common than systemic reactions in the adolescent cohort. After the first vaccination, the most common local reaction was pain at the injection site, 13.2%. The three most common systemic reactogenicities were fatigue (8.7%), myalgia (7.7%), and headache (7.7%). In contrast with the adolescent cohort, the local reactions were less reported than systemic reactions in the adult cohort. After the first vaccination, the three most common systemic reactogenicities were drowsiness (15.6%), myalgia (12.8%), and fatigue (11.6%). The local reaction was reported only 7.2%.

#### Adverse Events

Adverse events were reported approximately 3.4% in the adolescent cohort and 2% in the adult cohort. There was no vaccine-related adverse event reported in the adolescent cohort. On the other hand, vaccine-related adverse events were reported at 0.8% in the adult cohort. No severe, serious, life-threatening adverse event, death, or adverse event led to discontinuation reported in both cohorts (Table 2).

**Table 2.** Reporting of adverse events after dose 1 through 1 month after dose 2.

| Adverse event | 12-17 years old (N=234) | 18-30 years old (N=249) |
| --- | --- | --- |
|  | n (%) | n (%) |
| Any event | 8 (3.4%) | 5 (2%) |
| Related | 0 | 2 (0.8%) |
| Severe | 0 | 0 |
| Life-threatening | 0 | 0 |
| Any serious adverse event | 0 | 0 |
| Related |  |  |
| Severe |  |  |
| Life-threatening |  |  |
| Any adverse event leading to discontinuation | 0 | 0 |
| Related |  |  |
| Severe |  |  |
| Life-threatening |  |  |
| Death | 0 | 0 |

#### Immunogenicity

Four weeks after the second vaccination, the GMC of anti-RBD antibody in the adolescent cohort was 102.9 BAU/mL (95%CI; 91.0-116.4). The GMC of anti-RBD antibody in the adult cohort was 36.9 BAU/mL (95%CI; 30.9-44.0) (Figure 4). The GMR of the adolescent cohort was 2.79 (95%CI; 2.25-3.46, p-value; <0.0001) compared with the adult cohort (Table 3). The immune response of the adolescent cohort met non-inferiority criteria compared with the adult cohort (a lower limit of the two-side 95% CI > 0.67). Moreover, the lower limit of two-side 95% CI was more than 1, indicating the superiority of immune response in an adolescent cohort compared with the adult. Subgroup analysis also showed that the GMR of participants aged 12-14 years was 3.37 (95%CI; 2.59-4.38, p-value; <0.0001) and 2.27 (95%CI; 1.74-2.98, p-value; < 0.0001) in adolescents aged 15-17 years (Table 4).

**Figure 4.**
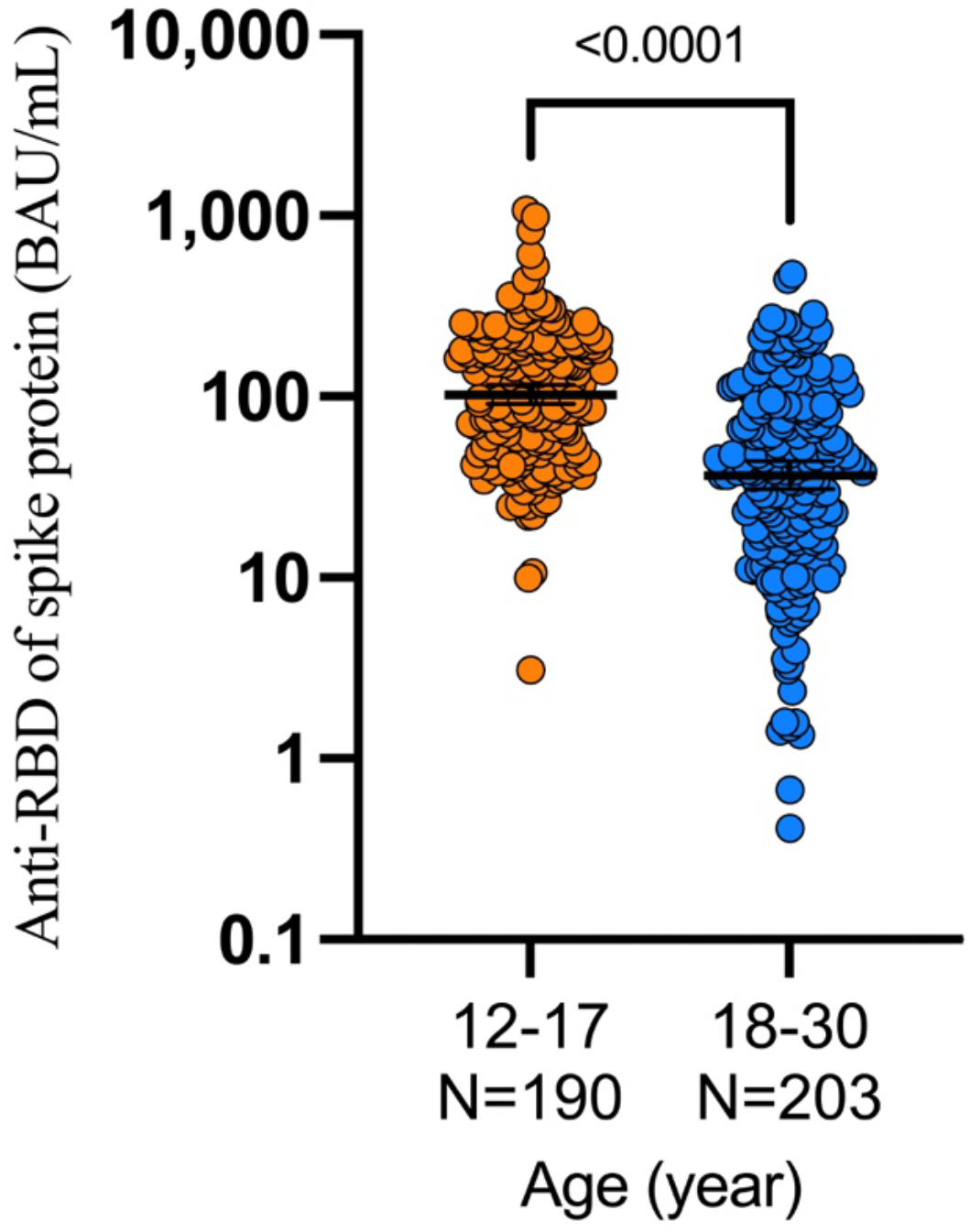
concentration of anti-receptor binding domain of spike protein of SARS-CoV-2. BAU: binding antibody unit. N: number of participants for immunogenicity analysis.

**Table 3.** Geometric mean concentration of each study group.

|  | 12-17 year | 18-30 year |
| --- | --- | --- |
| Number of participants | 190 | 203 |
| GMC (95%CI) (BAU/mL) | 102.9 (91.0-116.4) | 36.9 (30.9-44.0) |
| GMR (95%CI) | 2.79 (2.25-3.46) **** | reference |
| GMC; geometric mean concentration. GMR; Geometric mean ratio.<br>95%CI; 95% confidence interval. ****; p-value < 0.0001. BAU; binding antibody unit |  |  |

**Table 4.** Geometric mean concentration of each sub-group.

|  | 12-14 year | 15-17 year | 18-30 year |
| --- | --- | --- | --- |
| Number of participants | 99 | 91 | 203 |
| GMC (95%CI) (BAU/mL) | 124.2 (105.8-145.7) | 83.9 (69.8-100.7) | 36.9 (30.9-44.0) |
| GMR (95%CI) | 3.37 (2.59-4.38) **** | 2.27 (1.74-2.98) **** | reference |
| GMC; geometric mean concentration. GMR; Geometric mean ratio. 95%CI; 95% confidence interval. ****; p-value < 0.0001. BAU; binding antibody unit. |  |  |  |

## DISCUSSION

Two doses of the BBIBP-CorV vaccine with a 21-day interval can elicit a more robust immune response in adolescents aged 12-17 than adults aged 18-30 years at one month after the second dose with similar safety profiles. A robust immune response was observed in both younger (aged 12-14 years) and older (aged 15-17 years). The phase 3 trial of the BBIBP-CorV vaccine showed a high vaccine efficacy, 78.1%, in participants aged 18 years or more. (13) The result of our study inferred the vaccine effectiveness of the BBIBP-CorV vaccine in adolescents aged 12-17 years through the immunobridging approach. The safety profile after vaccination with the BBIBP-CorV vaccine in adolescents was slightly less than in adults, both reactogenicity and non-solicit adverse events.

Our study result is similar to the other study that evaluated the immunogenicity and safety of the adolescent in the other vaccine platform. The study of Robert W. Frenck et al. assessed the safety and immunogenicity of the BNT162b2 vaccine, mRNA vaccine platform, in adolescents aged 12-15 years compared with 16-25 years. The adverse events were comparable between participants ages 12-15 and 16-25 years. There was no vaccine-related serious adverse event in both cohorts. The immunogenicity was superior in participants ages 12-15 years than 16-25 years with GMR 1.76 (95%CI, 1.47-2.10). (10) Moreover, no participant received BNT162b2 developed Covid-19 7 days after the second vaccination. On the other hand, 16 of 978 participants received placebo developed Covid-19. Post-authorization vaccine effectiveness evaluations supported the immunobridging result of the BNT162b2 vaccine. The effectiveness of the BNT162b2 against hospitalization in participants aged 12-18 years was also evaluated using a test-negative design in 19 pediatric hospitals. The case group was the hospitalized patients with COVID-19-like symptoms with positive SARS-CoV-2 RT-PCR results. Two controlled groups were 1) hospitalized patients with COVID-19 like symptoms with negative SARS-CoV-2 RT-PCR or antigen test results (test-negative) and 2) hospitalized patients without COVID-19 associated symptoms. Six of 179 participants in case-patients were vaccinated with the BNT162b2. 93 of 285 in the controlled group received the BNT162b2. The vaccine effectiveness against hospitalization was 93% (95%CI, 83-97). (14) Similar results for safety and immunogenicity were demonstrated in participants aged 5-11 years after 2 doses of 10 μg of the BNT162b2 vaccine at 21 days intervals. One thousand five hundred seventeen children were randomized to receive the BNT162b2 vaccine, and 751 received a placebo. The safety profile was similar to the other age group. There were no vaccine-related serious events observed. The immunogenicity was non-inferior compared with the adult aged 16-25 years. The GMR of neutralizing titer in children aged 5-11 to those in 16-25 years was 1.04 (95%CI, 0.93-1.18), which met the non-inferiority criterion. (9)

The safety profile of our study did not differ from the phase 2 clinical trial of the BBIBP-CorV vaccine. The phase 1/2 clinical trial of the BBIBP-CorV vaccine aimed to evaluate the safety profile and immunogenicity in participants aged 3-17 years. The participants were stratified according to age (3-5 years, 6-12 years, and 13-17 years) and dose (2 μg, 4 μg, and 8 µg) group and randomized to receive vaccine or placebo. The vaccines were scheduled in three doses (days 0, 28, and 56). There were 288 participants in phase 1and 720 in the phase 2 trial. The adverse reactions were evaluated within 30 days after the whole vaccination procedure. The immunogenicity was assessed 28 days after dose 1, dose 2, and dose 3 of the BBIBP-CorV vaccine using neutralizing antibody titer. The adverse reactions were mild to moderate in severity in most cases. Pain at the injection site was the most common local reaction in participants aged 13-17 years, 7.9%. Fever was the most common systemic reaction, 10.3%. At the immunogenicity outcome, the immune responses of the 4 μg and 8 μg groups were higher than the 2 μg group. By days 28 after dose 2 vaccine in cohort 3-5 years, the GMT was 105.3 (95% CI; 95.4-116.2) in the 2 μg group, 180.2 (163.4-198.8) in the 4 μg group, and 170.8 (202.2-249.1) in the 8 μg group. The GMT ranged from 84.1 to 168.6 in the 6-12 years cohort and 88.0 to 155.7 in the 13-17 years cohort. (11)

To our knowledge, this is the first study that evaluated the effectiveness of the BBIBP-CorV vaccine using the immunobridging approach. The result of our study supported using the BBIBP-CorV vaccine in adolescents aged 12-17 years. Although most adolescents had a mild form of COVID-19 disease (1), vaccination in this group also had benefits. First of all, vaccination can prevent severe COVID-19 disease, especially in underlying comorbidity adolescents (14). Therefore, vaccination in this group should decrease morbidity and mortality. Second, evidence supports that vaccination with mRNA COVID-19 vaccine can prevent the multisystem inflammatory syndrome in children (MIS-C), a severe complication associated with SARS-CoV-2 infection in children and adolescents. At the end of December 2021, 76.7% of adolescents in France were vaccinated at least 1 dose of COVID-19 vaccine, mostly BNT162b2. In this period, there were 33 adolescents hospitalized due to MIS-C. Among them, 7 had received 1 dose of vaccine, and 26 were unvaccinated. No adolescent fully vaccinated with the COVID-19 vaccine developed MIS-C in this period. The hazard ratio for MIS-C in vaccinated participants was 0.09 (95%CI, 0.04-0.21). (3) Third, vaccination of adolescents can improve herd immunity for community prevention. (15)

Moreover, unlike the other vaccine platform where younger participants may develop severe adverse events, there were no special adverse events related to the BBIBP-CorV vaccine. For example, after vaccination with the mRNA162b2, a small proportion of younger participants developed myocarditis, especially the male gender. (16-18) The vaccine-induced thrombotic thrombocytopenia (VITT) is associated with the adenoviral vector platform, especially for the young population. (19)

This study had some limitations. First, we conducted a prospective cohort study of adolescents compared with our previous study of adult participants instead of a parallel clinical trial. Most adult participants were vaccinated when the study started. It was difficult to enroll the adult participants without prior vaccination with the COVID-19 vaccine. So, there is a difference in the case record form between the adolescent and adult cohorts. There is only a single questionnaire for local reaction in the adult cohort. However, the other solicited adverse events were the same. Second, we used the text message questionnaires instead of the diary book. Some participants in the adolescent cohort did not respond to the questionnaires about safety profiles. This limitation may improve by the active contact with the participants that do not complete the questionnaires.

In conclusion, vaccination with the BBIBP-CorV vaccine in the adolescent participants was safe and more robust immune response than adults aged 18-30 years.

## Data Availability

All data produced in the present work are contained in the manuscript.

## Acknowledgments

We thank the clinical research management unit for managing this project. We also thank the central laboratory of Chulabhorn Hospital for the laboratory testing. We gratefully acknowledge funding from Chulabhorn Royal Academy.

## Ethics

The study was conducted in accordance with the principles laid out in the Declaration of Helsinki guidelines for research involving human subjects. The study protocol was reviewed and approved by the Ethics Committee for Human Research, Chulabhorn Research Institute (Certificate No. 123/2564)

## References

1. Castagnoli R, Votto M, Licari A, et al. Severe Acute Respiratory Syndrome Coronavirus 2 (SARS-CoV-2) Infection in Children and Adolescents: A Systematic Review. JAMA Pediatr. Sep 1 2020;174(9):882–889. doi:10.1001/jamapediatrics.2020.1467

2. Preston LE, Chevinsky JR, Kompaniyets L, et al. Characteristics and Disease Severity of US Children and Adolescents Diagnosed With COVID-19. JAMA Netw Open. Apr 1 2021;4(4):e215298. doi:10.1001/jamanetworkopen.2021.5298

3. Levy M, Recher M, Hubert H, et al. Multisystem Inflammatory Syndrome in Children by COVID-19 Vaccination Status of Adolescents in France. JAMA. Dec 20 2021;doi:10.1001/jama.2021.23262

4. Feldstein LR, Tenforde MW, Friedman KG, et al. Characteristics and Outcomes of US Children and Adolescents With Multisystem Inflammatory Syndrome in Children (MIS-C) Compared With Severe Acute COVID-19. JAMA. Mar 16 2021;325(11):1074–1087. doi:10.1001/jama.2021.2091

5. Polack FP, Thomas SJ, Kitchin N, et al. Safety and Efficacy of the BNT162b2 mRNA Covid-19 Vaccine. N Engl J Med. Dec 31 2020;383(27):2603–2615. doi:10.1056/NEJMoa2034577

6. Baden LR, El Sahly HM, Essink B, et al. Efficacy and Safety of the mRNA-1273 SARS-CoV-2 Vaccine. N Engl J Med. Feb 4 2021;384(5):403–416. doi:10.1056/NEJMoa2035389

7. Falsey AR, Sobieszczyk ME, Hirsch I, et al. Phase 3 Safety and Efficacy of AZD1222 (ChAdOx1 nCoV-19) Covid-19 Vaccine. N Engl J Med. Sep 29 2021;doi:10.1056/NEJMoa2105290

8. Al Kaabi N, Zhang Y, Xia S, et al. Effect of 2 Inactivated SARS-CoV-2 Vaccines on Symptomatic COVID-19 Infection in Adults: A Randomized Clinical Trial. JAMA. May 26 2021;doi:10.1001/jama.2021.8565

9. Walter EB, Talaat KR, Sabharwal C, et al. Evaluation of the BNT162b2 Covid-19 Vaccine in Children 5 to 11 Years of Age. N Engl J Med. Jan 6 2022;386(1):35–46. doi:10.1056/NEJMoa2116298

10. Frenck RW, Jr., Klein NP, Kitchin N, et al. Safety, Immunogenicity, and Efficacy of the BNT162b2 Covid-19 Vaccine in Adolescents. N Engl J Med. Jul 15 2021;385(3):239–250. doi:10.1056/NEJMoa2107456

11. Xia S, Zhang Y, Wang Y, et al. Safety and immunogenicity of an inactivated COVID-19 vaccine, BBIBP-CorV, in people younger than 18 years: a randomised, double-blind, controlled, phase 1/2 trial. Lancet Infect Dis. Sep 15 2021;doi:10.1016/S1473-3099(21)00462-X

12. Resman Rus K, Korva M, Knap N, Avsic Zupanc T, Poljak M. Performance of the rapid high-throughput automated electrochemiluminescence immunoassay targeting total antibodies to the SARS-CoV-2 spike protein receptor binding domain in comparison to the neutralization assay. J Clin Virol. Jun 2021;139:104820. doi:10.1016/j.jcv.2021.104820

13. Al Kaabi N, Zhang Y, Xia S, et al. Effect of 2 Inactivated SARS-CoV-2 Vaccines on Symptomatic COVID-19 Infection in Adults: A Randomized Clinical Trial. JAMA. Jul 6 2021;326(1):35–45. doi:10.1001/jama.2021.8565

14. Olson SM, Newhams MM, Halasa NB, et al. Effectiveness of Pfizer-BioNTech mRNA Vaccination Against COVID-19 Hospitalization Among Persons Aged 12-18 Years - United States, June-September 2021. MMWR Morb Mortal Wkly Rep. Oct 22 2021;70(42):1483–1488. doi:10.15585/mmwr.mm7042e1

15. de Beer PAM, Van den Abbeele K. Inviting adolescents aged 12-17 for covid-19 vaccination: the need for patience. BMJ. Sep 9 2021;374:n2172. doi:10.1136/bmj.n2172

16. Mevorach D, Anis E, Cedar N, et al. Myocarditis after BNT162b2 mRNA Vaccine against Covid-19 in Israel. N Engl J Med. Dec 2 2021;385(23):2140–2149. doi:10.1056/NEJMoa2109730

17. Witberg G, Barda N, Hoss S, et al. Myocarditis after Covid-19 Vaccination in a Large Health Care Organization. N Engl J Med. Dec 2 2021;385(23):2132–2139. doi:10.1056/NEJMoa2110737

18. Larson KF, Ammirati E, Adler ED, et al. Myocarditis After BNT162b2 and mRNA-1273 Vaccination. Circulation. Aug 10 2021;144(6):506–508. doi:10.1161/CIRCULATIONAHA.121.055913

19. Pavord S, Scully M, Hunt BJ, et al. Clinical Features of Vaccine-Induced Immune Thrombocytopenia and Thrombosis. N Engl J Med. Oct 28 2021;385(18):1680–1689. doi:10.1056/NEJMoa2109908

